# Perceptions and Misconceptions of PSA Screening in Switzerland: A Preference Epidemiology Study

**DOI:** 10.1101/2024.11.06.24316816

**Authors:** Giovanni Spitale, Federico Germani, Nikola Biller-Andorno

## Abstract

**Introduction:** PSA screening for prostate cancer detection is highly debated due to the challenging balance between its potential benefits and risks, regarding overdiagnosis and overtreatment. This study applies a preference epidemiology approach to understand how individuals evaluate these trade-offs, aiming to identify the thresholds at which people find screening acceptable or burdensome. By examining both personal and societal perspectives on PSA screening, this preference epidemiology study provides insights into how values, preferences, and psychosocial factors influence health-related decision-making.

**Methods:** A survey of Swiss men aged 55+ examined their awareness of PSA screening, their screening history, and their willingness to participate in future screenings. Hypothetical scenarios illustrating different trade-offs between overdiagnosis and lives saved by PSA screening were presented to the participants. Data were analyzed using Chi-square tests, MANOVA, and thematic analysis.

**Results:** 425 participants were included in the study. Most respondents significantly overestimated PSA screening’s life-saving potential, with a median estimate of 50 deaths prevented versus the current figure of 3 deaths prevented per 1000 persons screened reported in the literature. Over half of the participants supported the use of PSA screening even in a hypothetical scenario where no lives were saved. Personal and family cancer history were associated with increased support for PSA screening.

**Discussion and conclusion:** Providing factual information about the risks and benefits of PSA screening alone may not ensure fully informed, autonomous decision-making. A systematic understanding of how personal evaluations of the risks and benefits are conducted is essential for the assessment of screening programs, which could inform key policy decisions, such as the integration of screenings into mandatory health insurance packages. These findings highlight the importance for both policy decisions and health communication to go beyond fact-sharing and incorporate systematic evidence from nuanced, value-sensitive evaluations to better support informed and autonomous decision-making.

## Introduction

Preference epidemiology is an emerging field that explores how individuals’ values, preferences, and perceptions shape their health-related decisions, particularly when faced with complex trade-offs between benefits and harms. Preference epidemiology aims to study how people weigh options in various healthcare scenarios, where they must consider complex trade-offs between benefits and risks. It examines the thresholds at which individuals are willing to accept certain risks, such as overdiagnosis or overtreatment, in exchange for perceived benefits like early detection. By identifying these thresholds for acceptability, preference epidemiology provides valuable data that can inform both public and health policy debates, helping to create guidelines that align with patient values.

The ongoing debate surrounding mammography screening for women in their 40s underscores the importance of preference epidemiology [1]. In 2002, the United States Preventive Services Task Force (USPSTF) gave a “B” grade recommendation, advocating routine screening mammography for women in this age group based on perceived benefits [1]. However, seven years later, this recommendation was downgraded to a “C” grade, not against screening but suggesting that decisions should be made individually through discussion with healthcare providers, considering the delicate balance of benefits and harms [1]. This shift towards a nuanced recommendation was met with public confusion and criticism. While assigning this grade did not imply that mammography should not be available to women in this age group, it highlighted the need for women to weigh the complex balance of benefits and risks with their healthcare providers and make a personal decision. Despite the USPSTF accurately noted it did not recommend “against routine screening” for women aged 40-49, many interpreted this shift as a move opposing mammography altogether [1]. Critics of this choice argued that the new grade placed women at risk. The task force’s stance did not change, but the controversy highlighted the challenge of communicating complex risk-benefit analyses to the public, especially when values and perceptions vary widely [1]. In its most recent 2024 recommendation [2,3], the USPSTF again adjusted the recommendation of routine screening mammography to a “B” grade, partially based on decision modeling [4]. This statistical approach suggested that starting screenings at age 40 could prevent an additional 1.3 breast cancer deaths per 1,000 women, with a potentially greater benefit among Black women, who experience higher breast cancer mortality rates. This decision again reflects a different interpretation of the balance between benefits and harms.

Another aim of preference epidemiology is to foster open communication about the burdens and benefits of medical interventions, encouraging a transparent dialogue that supports health literacy and informed decision-making. This is important because, as we learn more about how specific populations evaluate trade-offs, we can better tailor healthcare communication and interventions to meet their needs, ultimately enhancing informed decision making and autonomy. Additionally, insights gained from preference epidemiology can aid in resource allocation by identifying which interventions are most acceptable and effective for a given population. The more we understand how people think about these complex choices, the better equipped we are to design public health programs that are both patient-centered and ethically sound. Preference epidemiology plays a crucial role in such decisions by examining how different populations respond to risks and benefits, providing a systematic understanding that supports clearer and more personalized guidance. By capturing these complex trade-offs, preference epidemiology enhances public health efforts, ensuring that recommendations resonate with individuals’ values and support informed, autonomous decision-making. Insights from this field can also inform resource allocation, identifying the most acceptable and effective interventions for different populations, ultimately contributing to patient-centered, ethically sound healthcare policies.

In this study, prostate-specific antigen (PSA) screening will serve as a test case for the preference epidemiology approach. PSA screening is a blood test aimed at detecting prostate cancer in its early stages [5]. It has been a subject of significant debate within the medical community due to the balance between its potential benefits and burdens [6,7]. While PSA testing can detect prostate cancer before it causes symptoms, potentially improving recovery chances and lowering the risk of metastasis [7], the benefits of routine PSA screening for reducing prostate cancer-related deaths have not been convincingly demonstrated [8], in consideration of the trade-off with the potential burdens related to overdiagnosis and overtreatment [9,10]. As a result, organizations such as the Canadian Task Force on Preventive Health Care and the US Preventive Services Task Force do not recommend including PSA testing in periodic health examinations for men of any age [8], and the German Institute for Quality and Efficiency in Health Care (IQWiG) recognizes that the benefit of population-based PSA screening for men with an average risk of prostate cancer does not outweigh the harm caused [11,12]. These considerations on harm reduction have been also brought forth by the European Randomised study of Screening for Prostate Cancer (ERSPC) in 2014, which recommended considering the clarification of the potential harms (overdiagnosis and overtreatment) as “a prerequisite for the introduction of population-based screening” [13]. Despite this, PSA testing continues to be widely used [8]. The decision for men to undergo PSA screening is complex and influenced by a range of factors, such as individual preferences, values, and decision-making processes that can affect health outcomes. In this study, we explore these factors with a ‘preference epidemiology’ approach.

Research has shown that men generally prefer an informed and transparent approach to PSA screening rather than covert testing [14]. Understanding and recognizing these preferences is essential for developing public health policies that are acceptable to the target population [15]. It also emphasizes the importance of shared decision-making between physicians and patients, considering individual risk factors, preferences, and values to guide appropriate screening choices [16].

One of the primary trade-offs men consider when deciding about PSA screening is the balance between the benefits of early detection and the risks of overdiagnosis and overtreatment. Overdiagnosis refers to the detection of cellular changes or tumors through screening that would not have caused any problems if left undetected [10,12]. These cases may lead to unnecessary treatment, potentially causing harm [7,9]. In 2014 the European Randomised study of Screening for Prostate Cancer (ERSPC) has shown that the absolute risk reduction in prostate cancer mortality over 13 years with screening intervals of 4 years (2 in Sweden) was 0.11 per 1000 person-years, or 1.28 per 1000 men randomized, meaning that one prostate cancer death was prevented for every 781 men screened [13]. More recently, the IQWiG report on PSA screening, published in 2020 and based on the analysis of 11 randomized controlled trials with more than 400,000 participants worldwide, concluded that PSA screening could prevent about 3 prostate cancer deaths per 1000 men screened over 16 years. However, overdiagnosis can occur in up to 60 out of 1,000 men participating in such a screening program [7,11]. These results contributed to fuel the decade-long controversy on PSA screening [17,18]. It is worth noting that newer, more specific, diagnostic tests [19] along with imaging approaches [20] may help reduce the risks of overdiagnosis and overtreatment. As science progresses and follow-up testing becomes more precise, allowing for more targeted treatments, we can expect the benefit-burden ratio to improve over time. In this sense, current evidence reflects “old” practices. While we acknowledge the potential for future improvement in figures related to screening outcomes, our primary focus here is conceptual, and related to preference epidemiology. Specifically, we emphasize the need for preference epidemiology to capture and understand how individuals weigh the complex trade-offs involved in medical screening, with currently used screening technology. The framework remains relevant even as medical advancements change the picture, because the question of where people’s decision-making tipping point lies—based on the evaluation of the least acceptable benefit for a given burden, or vice versa—remains. Moving forward, public health recommendations and medical information will, of course, be updated to reflect new evidence, but the core issue of how people make decisions about screening remains critical to address, regardless of advances in screening technology and individual diagnostic follow-up tests.

Research has shown that most men believe PSA testing is beneficial and can significantly reduce the risk of dying from prostate cancer [8,10]. However, there is limited data on patient understanding of the risks associated with overdiagnosis and overtreatment [8]. To make informed decisions, men need to be fully informed about both the benefits and potential downsides of PSA testing [15]. Studies suggest that informed deliberation of the harms and benefits of PSA screening can influence men’s screening decisions [15]. Several factors can influence an individual’s decision to undergo PSA screening, including having a family history of prostate cancer: men with a family history of prostate cancer may be more likely to choose screening due to fear of the disease [16]; recommendations from family and friends: the influence of wives and other family members can play a significant role in men’s decisions to pursue screening [16]; physician recommendations and communication: while physician discussions are highly predictive of screening behavior, the quality and content of these conversations vary [16]. Some men may not receive adequate information about the risks associated with PSA screening, while others may already be committed to screening before discussing it with their doctor [16,21].

This study explores the psychosocial factors influencing men’s decisions regarding PSA screening for prostate cancer, within the framework of preference epidemiology. The goal is to understand how individuals weigh the benefits of early detection against the risks of overdiagnosis and overtreatment— central issues in the PSA screening debate, and whether their perception of benefits and burden is aligned with evidence. Despite recommendations to limit PSA testing due to its benefit/harm ratio, the procedure remains widely used, highlighting a need to understand the personal and social factors driving these decisions. This study seeks to capture both individual thresholds for acceptability and societal perceptions of PSA screening’s value. These insights contribute to understanding how informed decision-making shapes attitudes towards screening, offering valuable information for developing more patient-centered public health policies and screening strategies.

## Methods

This cross-sectional study aims to assess the psychosocial factors influencing decisions regarding PSA screening for prostate cancer, as well as the public perception in terms of lives saved and the overall perceived value of PSA screening programs of Swiss males aged 55 and above. The survey through which the data for this study were collected explores participants’ knowledge of PSA screening, family history of prostate cancer, and their perceived risk of prostate cancer, as well as their willingness to undergo screening, and the perceived societal value of PSA screening programs. The survey was made available in German, French, Italian, and English. The survey structure and data are available on this study’s OSF repository [22].

### Survey structure

The survey was designed using a structured questionnaire, which was hosted on PubliCo [23,24], a secure platform developed and owned by the University of Zurich. The first section of the survey explains the purpose of the study, explaining the voluntary nature of participation and the anonymity of all responses. This is followed by a series of demographic questions that collect basic information such as age, education level, and family history of prostate cancer [25]. Participants’ level of worry about prostate cancer [26] and understanding of PSA screening were assessed. The survey also explores participants’ past and current screening behaviors, including whether they had previously undergone a PSA test and their reasons for either participating or not in future screenings [26]. Participants were then provided with detailed information about the PSA test procedure. This includes an explanation of how the test works, its purpose in detecting prostate cancer, and the potential outcomes. Participants were then presented with a question aimed at assessing their understanding of the test’s benefit from a population-level perspective. Specifically, they were asked: *“How many deaths from prostate cancer can be prevented by prostate screening if 1,000 people are screened once every 4 years over a period of 16 years?”* This question is designed to evaluate participants’ baseline knowledge about the potential life-saving benefits of PSA screening, drawing on data from previous large-scale studies [11,13]. By posing this question, we can gauge participants’ perceptions of the test’s population-level benefit before providing the actual figure and engaging them in the case assessment, where they must weigh the PSA screening’s trade-offs between lives saved and overdiagnosis. The trade-off between the benefits and burdens are presented both numerically and with an infographic **(Figure 1)**.

**Figure 1.**
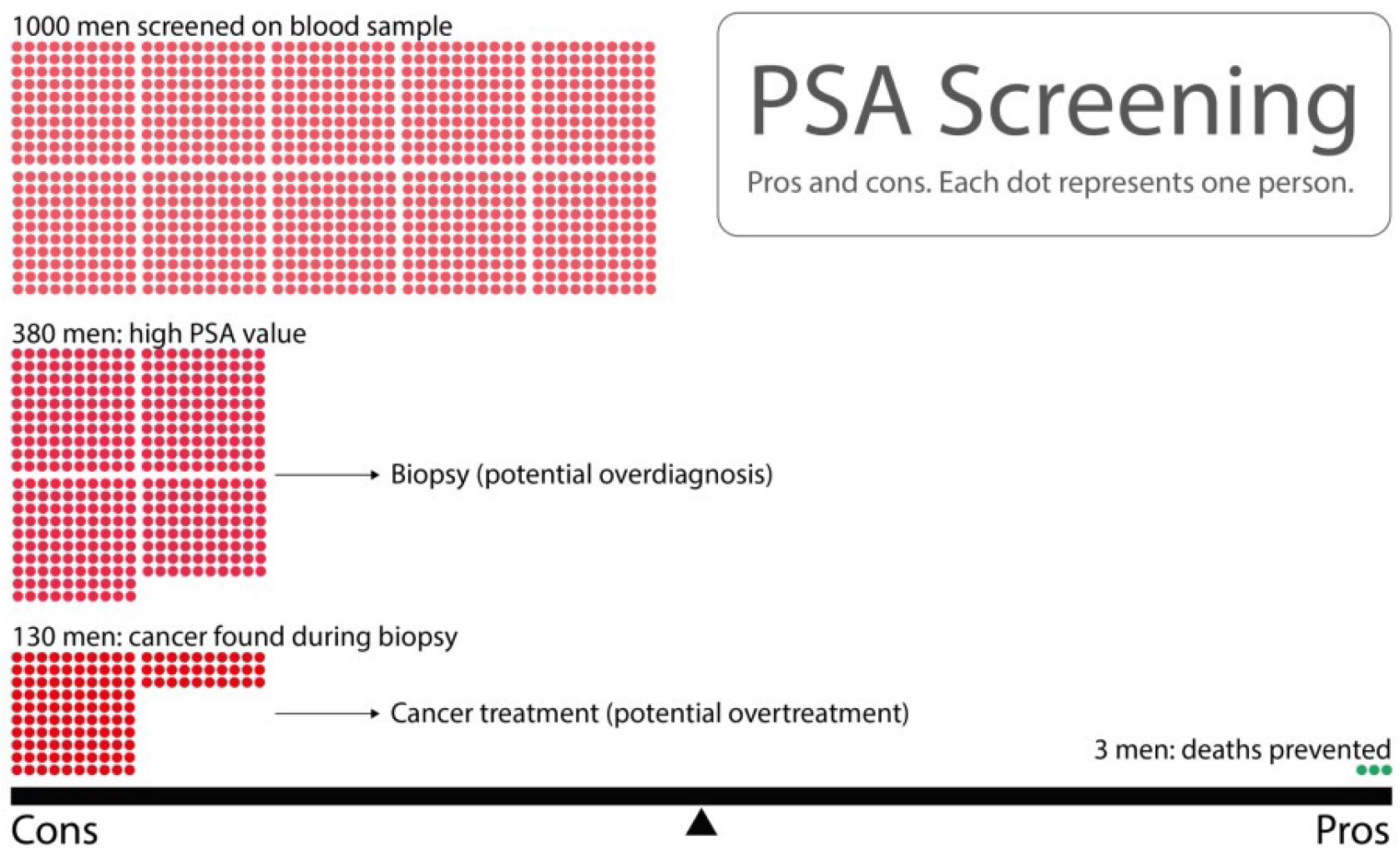
Trade-off between the pros and cons of PSA screening for prostate cancer. Out of 1,000 men screened, 380 may have elevated PSA levels, leading to biopsies, which may result in overdiagnosis. Of these, cancer may be detected in 130 men, potentially leading to overtreatment. PSA screening may prevent death in 3 men, showing the balance between the benefits (lives saved) and the burdens (overdiagnosis and overtreatment). Each dot represents one person, highlighting the scale of potential outcomes visually [7,12,13].

Participants’ attitudes toward PSA screening are then assessed through questions aimed at understanding both their willingness to undergo the test and the factors contributing to hesitancy. For those participants who indicate that they are either “Somewhat unwilling” or “Very unwilling” to undergo screening, a conditional follow-up question on reasons for hesitancy was presented.

In the “Case Assessment” section of the survey, participants were presented with hypothetical scenarios that illustrate the trade-offs between the risks of overdiagnosis or overtreatment and the potential number of lives saved by PSA screening. Initially, all participants were asked to assess a control case, referred to as Case 0, where the ratio of harm to benefit corresponds to 60 cases of overdiagnosis or overtreatment for 0 lives saved, representing an unrealistic scenario with no discernible benefit to screening. Participants who indicated they were not willing to undergo PSA screening in Case 0 were also asked: “What number of deaths prevented would make the outcomes acceptable to you, considering the balance with overdiagnosis and overtreatment?” We used their responses to identify the “tipping point” for each participant—i.e., the minimum benefit they would require for considering the use of PSA screening acceptable. Following the control case, participants were randomly assigned to assess one additional hypothetical case from a range of scenarios, each presenting a fixed number of 60 cases of overdiagnosis or overtreatment, balanced against varying numbers of lives saved. Cases are presented in **Table 1;** an example of the vignettes used is presented in **Figure 2**.

**Table 1.** Overview of the hypothetical PSA screening scenarios presented to participants. Each scenario maintains a constant number of 60 cases of overdiagnosis or overtreatment (harms), while varying the number of lives saved (benefits). The cases range from no benefit (60 vs 0) to substantial benefits (60 vs 500+), providing participants with a spectrum of trade-offs to evaluate in their decision-making process.

| Case | Number of cases of Overdiagnosis/Overtreatment (Harms) vs Number of Lives Saved (Benefits) |
| --- | --- |
| Case 0 | 60 vs 0 (no benefit) |
| Case 1 | 60 vs 1-2 |
| Case 2 | 60 vs 3 (actual benefit based on clinical data) |
| Case 3 | 60 vs 4-10 |
| Case 4 | 60 vs 11-20 |
| Case 5 | 60 vs 21-59 |
| Case 6 | 60 vs 60 |
| Case 7 | 60 vs 61-100 |
| Case 8 | 60 vs 101-500 |
| Case 9 | 60 vs 500+ |

**Figure 2.**
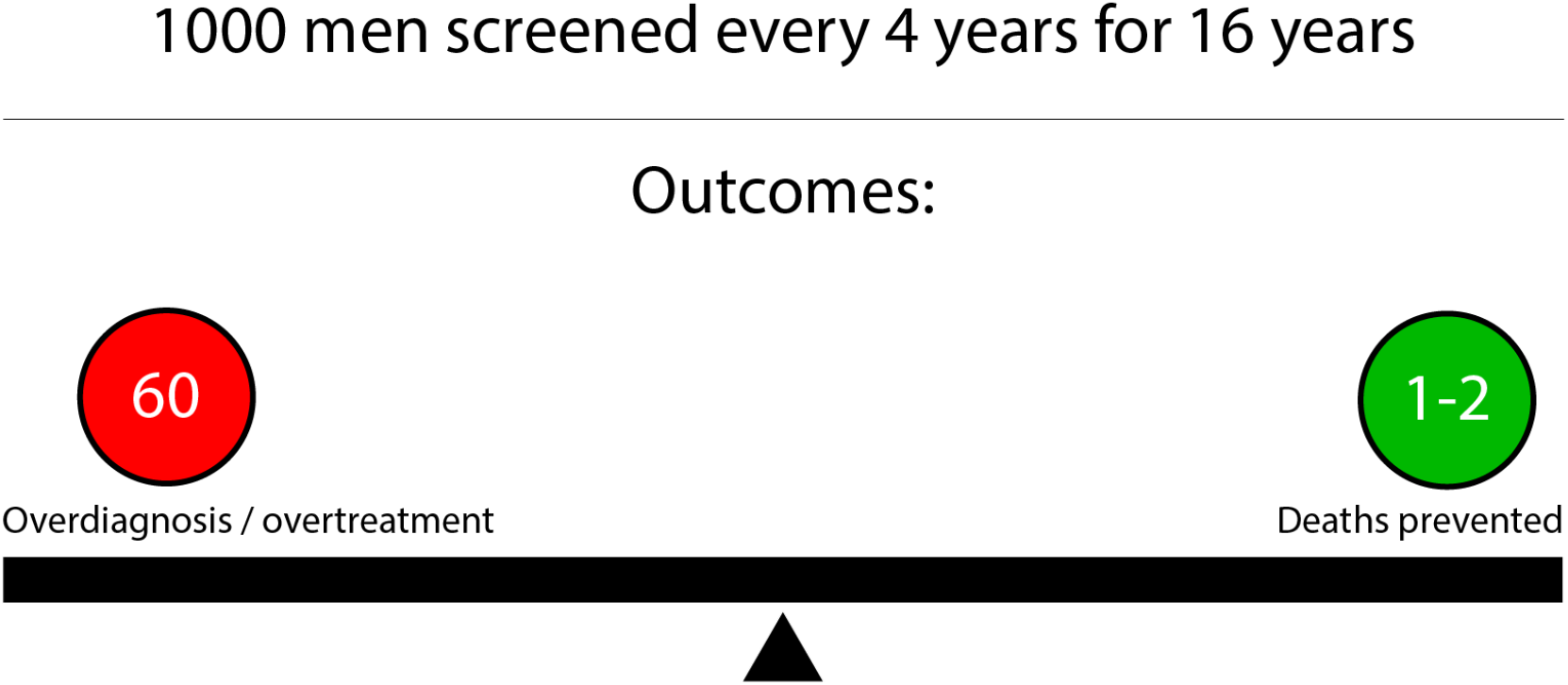
Example vignette used in the study, illustrating Scenario 1-2. 1,000 men are screened every 4 years for 16 years; the outcomes show 60 cases of overdiagnosis or overtreatment, with 1-2 deaths prevented.

Participants were asked to consider these scenarios and respond to the question: *“Consider this* ***hypothetical*** *scenario. Would you be willing to take a PSA test with this ratio of benefits to negative effects?”* We chose this approach of presenting fixed hypothetical scenarios rather than asking participants directly for their threshold because this method helps participants as they may find it difficult to accurately determine or articulate their own thresholds without being prompted by specific contextual examples. Following the case assessment, participants were presented with a final question designed to shift their focus from personal decision-making to a broader societal perspective, with participants expressing their views on the societal value of PSA screening programs and the value of PSA screening as a public health measure.

### Recruitment

To recruit participants for the study, two Facebook Ads campaigns were conducted, targeting Swiss males aged 55 and above across four language groups: English, German, French, and Italian. Details about the recruitment can be found on this study’s OSF repository [22].

### Statistical tests

In descriptive statistics outliers were identified and removed using the interquartile range (IQR) * 1.5 method [27]. We employed Chi-square tests to assess relationships between categorical variables, and Shapiro-Wilk tests to check for normality in continuous data. For non-normally distributed data, we applied the Mann-Whitney U test to compare groups. We conducted a multivariate analysis of variance (MANOVA) to assess the impact of demographic and health-related factors on two dependent variables: willingness to undergo PSA screening and perceived societal value of PSA screening. For significant MANOVA results, we performed analysis of variance (ANOVA) to further examine the role of specific independent variables on these outcomes. Post-hoc pairwise comparisons were conducted using Tukey’s Honest Significant Difference (HSD) test to identify specific group differences within the significant independent variables. All statistical analyses were performed in a Jupyter Notebook, utilizing the following Python libraries: pandas for data manipulation, matplotlib and seaborn for data visualization, and numpy for numerical computations. For statistical tests, we used scipy for Chi-square tests, statsmodels for conducting the MANOVA and ANOVA analyses, and pairwise_tukeyhsd from statsmodels for Tukey’s post-hoc tests. The code is available on this study’s OSF repository [22].

### Qualitative analysis

We conducted a thematic analysis [28] on the open-ended responses provided by participants who responded that they would be willing to undergo PSA screening with the hypothetical benefit/harm ratio presented by Scenario 0. After collecting their answers, we manually coded the text, identifying recurring themes that reflected the participants’ motivations. The themes were derived through an iterative process, where initial categories were established based on the content of the responses. These categories were then refined and consolidated as more responses were reviewed. Each response could be assigned to one or more themes, depending on the content.

## Results

The data was cleaned by removing 83 responses where the survey duration was under 3 minutes, 1 response from a female participant, and 19 responses from participants with less than 46 years of age. After this process, the study is based on the remaining 425 responses. The majority of participants (87,44%) were Swiss nationals, with 47 participants (11,14%) from other nationalities. The age distribution was concentrated in the older age groups, with 180 participants (42,35%) aged 66-75, 132 participants (31,06%) aged 56-65, and 80 participants (18,82%) aged 76 and above. Only 33 participants (7,76%) fell into the 46-55 age range.

The majority of participants, 266 (63.03%), reported having undergone PSA screening within the past year, while an additional 87 participants (20.62%) had undergone the test more than a year ago. A total of 61 participants (14.45%) indicated they had never undergone PSA screening, and 7 participants (1.66%) were unsure about their screening history. Regarding personal cancer diagnosis, 295 participants (71.43%) reported never having been diagnosed with cancer, while 98 participants (23.73%) had been diagnosed with prostate cancer. An additional 19 participants (4.60%) indicated a diagnosis of another type of cancer. In terms of familiarity with prostate cancer, 317 participants (76.39%) reported no family history of prostate cancer, while 77 participants (18.55%) had at least one relative diagnosed with the disease. Familiarity with other types of cancer was slightly higher, with 214 participants (50.95%) reporting that they had one or more relatives diagnosed with cancer, including 148 participants (35.24%) with a single affected relative and 39 participants (9.29%) with two relatives affected. Complete demographic details are available in the OSF repository [22].

### Discrepancy between reality and estimations about PSA screening programs

#### Estimated deaths prevented far exceed actual deaths prevented

In the survey, participants were asked to estimate how many deaths could be prevented by PSA screening programs, with the actual figure from the literature being about 3 deaths per 1,000 men screened [11,13]. The responses from participants show a significant overestimation. After removing outliers, the average estimated number of deaths prevented was 196. The median estimate was 50 deaths. Results are illustrated in **Figure 3**.

**Figure 3.**
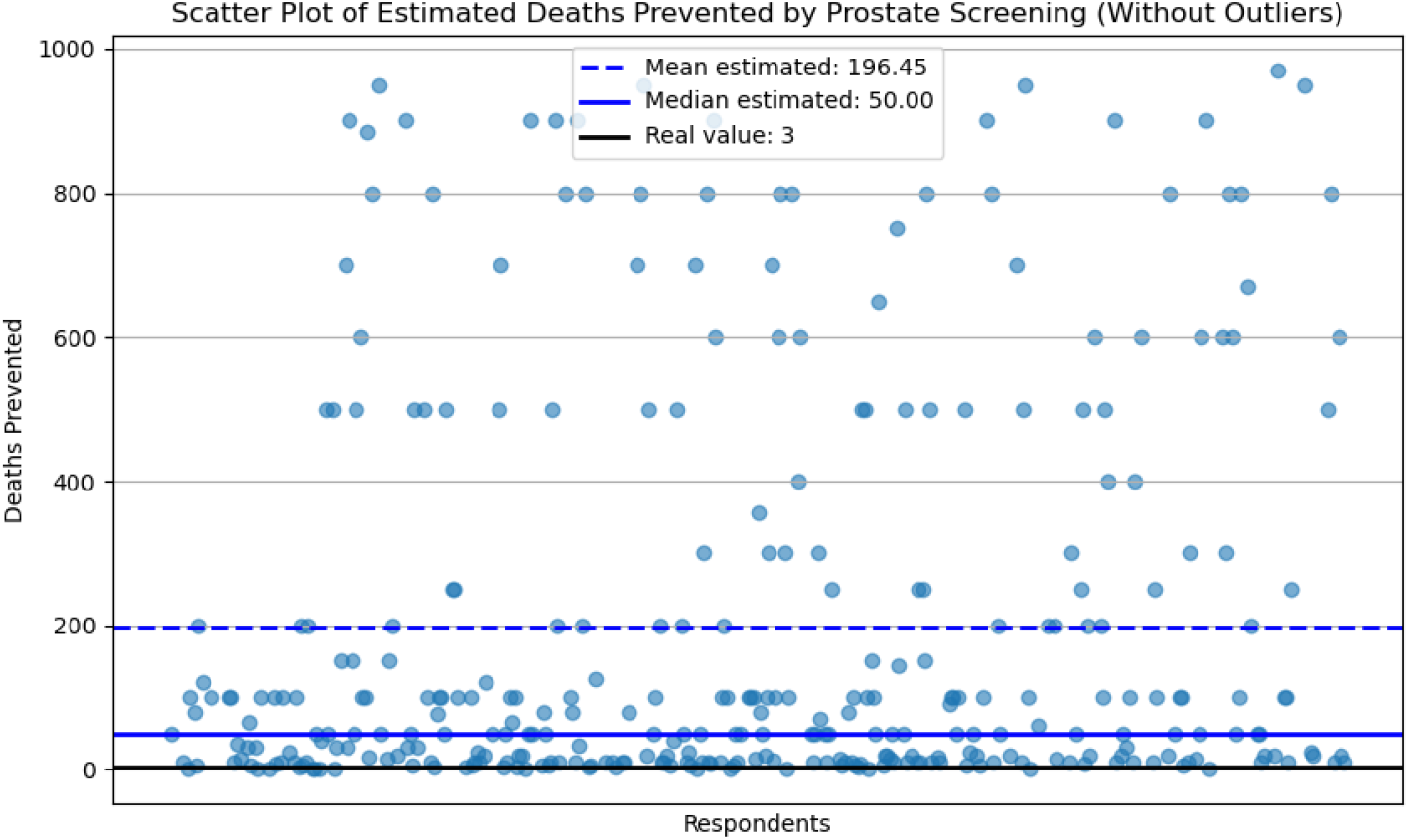
Scatter plot showing participants’ estimates of deaths prevented by prostate screening, after removing outliers. The blue dashed line represents the mean estimate of 196 deaths; the solid blue line represents the median estimate of 50 deaths prevented; while the black line indicates the real value from the literature, which is 3 deaths prevented per 1,000 men screened. Outliers were identified and removed using the interquartile range (IQR) * 1.5 method.

This discrepancy highlights a significant gap in public understanding of the actual benefits of PSA screening, as participants consistently overestimated the number of lives saved compared to the actual figure.

#### Participants consider PSA screening acceptable if it prevents a greater number of deaths than the actual number it prevents

Participants who were presented with Scenario 0 (60 cases of overdiagnosis/overtreatment versus no benefit in terms of deaths prevented) and answered “no” to participating in a PSA screening program with this risk benefit profile were asked a follow-up question: *“What number of deaths prevented (the green number) would make the outcomes acceptable to you, considering the balance with overdiagnosis/overtreatment?”*. As above, responses were cleaned to remove outliers. The descriptive statistics of the remaining responses showed that participants on average considered PSA screening acceptable when it led to preventing 30 deaths, which is ten times higher than the actual figure of 3 deaths prevented by PSA screening programs reported in the literature. The median estimate was of 19 deaths prevented, while the minimum and maximum acceptable estimates ranged from 1 to 100 deaths. Results are illustrated in **Figure 4**.

**Figure 4.**
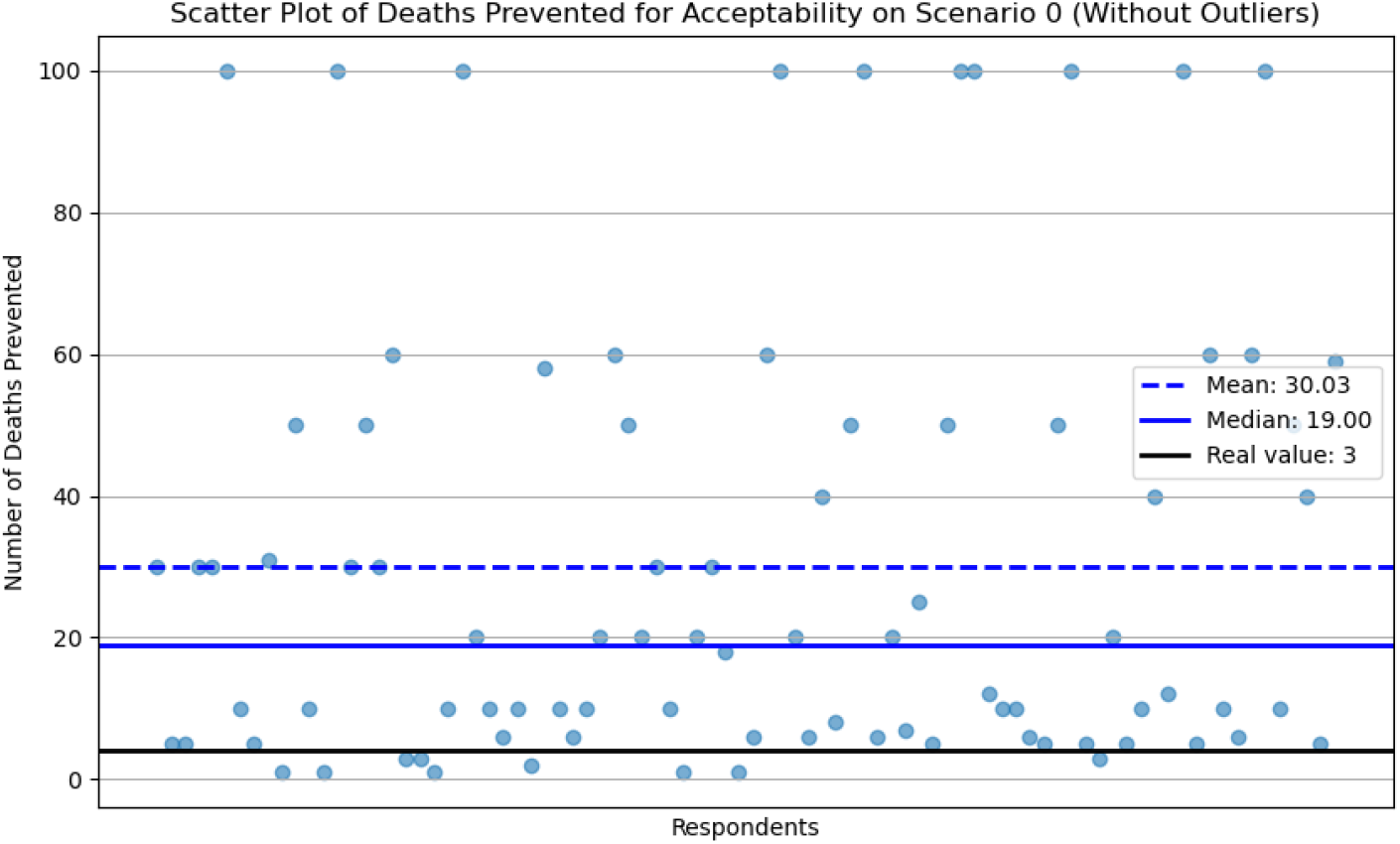
Scatter plot of estimated deaths prevented required for participants to find PSA screening acceptable in Scenario 0 (“tipping point”), after removing outliers. The blue dashed line indicates the mean estimate of 30 deaths prevented; the solid blue line represents the median estimate of 19 deaths prevented; while the black line indicates the real value from the literature, which is 3 deaths prevented per 1,000 men screened. Outliers were identified and removed using the interquartile range (IQR) * 1.5 method.

#### Supporting PSA screening in a hypothetical scenario where it offers no benefits is associated with an inflated perception of its advantages

Surprisingly, despite Scenario 0 was presented as a hypothetical scenario in which PSA screening offers no benefit, more than half of the participants (n= 143; 55.2%) still deemed the screening acceptable also if it caused overdiagnosis in 60 cases, and prevented no death; only 116 participants (44.8%) found PSA screening in Scenario 0 as an unnecessary measure.

In the analysis we compared the estimates of deaths prevented by PSA screening between participants who accepted Scenario 0 (“Yes” group) and those who did not (“No” group). Participants who deemed Scenario 0 acceptable, despite no lives being saved by PSA screening, estimated a significantly higher average number of deaths prevented by PSA screening (mean = 166, median = 80) compared with those who found Scenario 0 unacceptable (mean = 85, median = 20). Those willing to accept the screening program even when presented with Scenario 0 tend to overestimate the potential life-saving benefits of PSA screening more than those who reject it, indicating that inflated expectations and unconditional support for the benefits of PSA screening may play a role in their decision-making. Results are illustrated in **Figure 5**.

**Figure 5.**
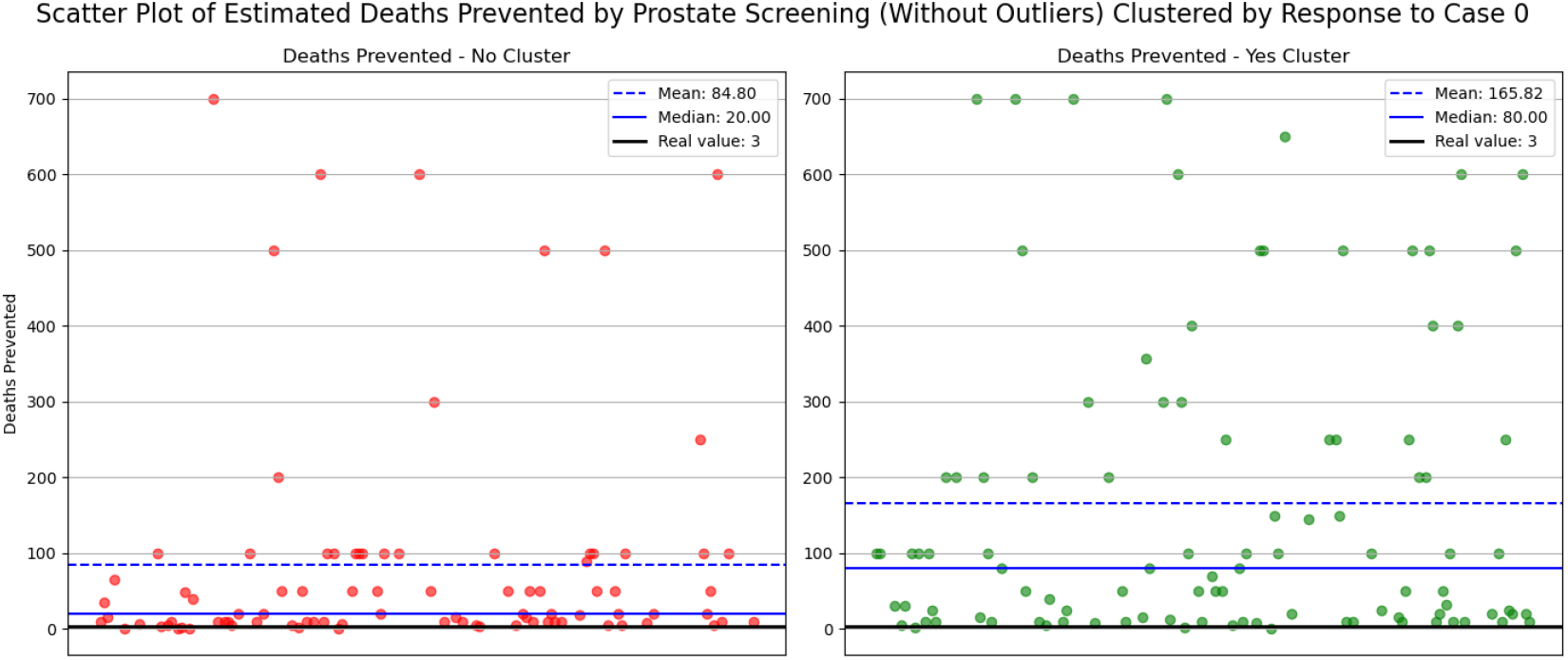
Scatter plot of estimated deaths prevented by PSA screening (without outliers), clustered by participants’ responses to Scenario 0. The left plot (“No Cluster”) shows the estimated number of deaths prevented by PSA screening from participants who rejected Scenario 0, while the right plot (“Yes Cluster”) shows estimates from participants who accepted Scenario 0. Dashed blue lines represent means; solid blue lines represent medians; solid black lines represent the actual number of deaths prevented by PSA screening from the literature. Outliers were identified and removed using the interquartile range (IQR) * 1.5 method.

In the comparison of estimated deaths prevented by PSA screening between participants who accepted Scenario 0 (i.e., “Yes” group) and those who rejected it (i.e., “No” group), normality tests were conducted using the Shapiro-Wilk test. The p-values for both the “Yes” group and the “No” group were < 0.001, indicating that the data in both groups did not follow a normal distribution. Consequently, the non-parametric Mann-Whitney U test was applied to compare the two groups. The test revealed a statistically significant difference between the “Yes” and “No” groups, with a p-value of 0.00036 (U = 5839.0).

#### Variation in acceptability of PSA screening across different hypothetical benefit scenarios

The acceptability of PSA screening across other scenarios (excluding Scenario 0) included in the study is fully illustrated in the contingency table and in the breakdown available in the study’s repository [22]. As described in **Figure 6**, as expected the overall acceptability was highest for scenarios with larger benefits, starting with the scenario in which 21 to 59 deaths could be prevented by PSA screening, where 97.6% of participants supported the use of PSA screening, with just 2.4% of respondents rejecting it. Scenarios with smaller benefits, such as 1-2 deaths prevented for 1000 men screened, resulted in lower acceptability, with only 80% of participants deeming it acceptable and 20% rejecting it. Scenario 3, which represents the actual risk/benefit profile of PSA screening based on clinical data (i.e. 3 deaths prevented for 1000 men screened), showed the highest rejection rate among participants. While these results show some general trends, such as increasing acceptability with greater perceived benefits until the scenario in which 60 deaths are prevented for 1000 men screened, the overall pattern is difficult to interpret, with varying support for PSA screening across the different scenarios.

**Figure 6.**
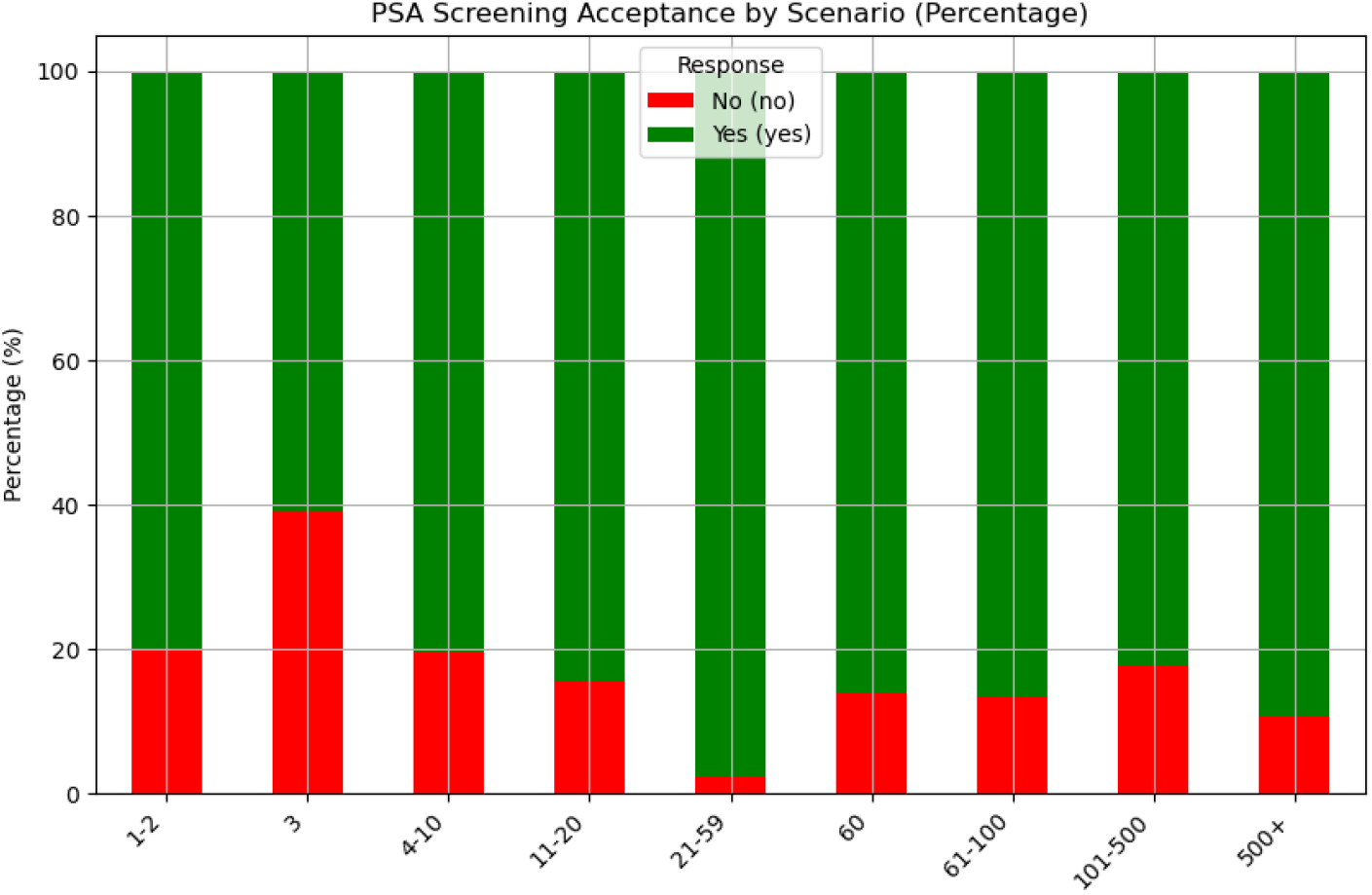
PSA screening acceptance by scenario (percentage). This bar chart displays the percentage of participants who found PSA screening acceptable (green) versus unacceptable (red) when presented with different scenarios. The numbers of the x axis indicate how many deaths were presented in the hypothetical scenario by PSA screening.

#### People maintain a positive opinion on the societal value of PSA screening programs

The final question in the survey asked participants to assess the overall societal value of PSA screening, after they had been informed about the risk/benefit profile, including the potential for overdiagnosis, overtreatment, and the actual number of deaths prevented (**Figure 1**). Despite this awareness, most respondents maintained a very positive view of PSA screening. A majority of 236 participants (55.5%) rated PSA screening programs as highly valuable, and 121 participants (28.5%) considered it moderately valuable. Only a smaller group expressed more critical views, with 51 participants (12%) stating that PSA screening was of limited value, and just 6 participants (1.4%) deeming it not valuable at all. These findings indicate that when presented with evidence about the benefits/risk profile of PSA screening, participants largely upheld their support for PSA screening and belief in its societal importance.

### Who are the PSA screening supporters?

#### Supporters of PSA screening are willing to partake PSA screening programs and believe they hold significant societal value, regardless of PSA screening’s benefits or lack thereof

The data reveals the existence of a specific group of PSA screening supporters who consistently overestimate the benefits of PSA screening and rejects the accuracy and truthfulness of the actual benefit/risk profile for PSA screening we provided in the survey. To better understand which factors may influence this behavior, we conducted a Chi Square test considering a comprehensive range of possible variables, including demographics, personal health history, attitudes towards PSA screening and its societal value, as independent variables possibly influencing the response to Scenario 0 (0 deaths prevented for 1000 men screened). Among these variables, two showed strong associations with participants’ willingness to accept Scenario 0: the willingness to undergo PSA screening (p < 0.0001), and the perceived societal value of PSA screening (p < 0.0001) (full data available in the study’s OSF repository [22]). We further explored the differences across different levels of these variables using post-hoc Chi Square tests (results and methodological details are available in the study’s OSF repository [22]), adjusting p-values with the Bonferroni correction. We find that the difference between those who are *very willing* to undergo PSA screening and those who are *somewhat unwilling* in their support for PSA screening in the context of Scenario 0 is highly significant (p < 0.0001, adjusted p < 0.0001); Even among participants who were generally positive about PSA screening, we find a noticeable distinction between those who are *very willing* and those that are *somewhat willing* to support PSA screening in the context of Scenario 0 (p = 0.003, adjusted p = 0.045).

The comparison of perceived societal value also highlights significant differences between participants. Those who rated PSA screening as *highly valuable* were significantly more likely to differ in their support for PSA screening in the context of Scenario 0 from those who rated it as *moderately valuable* (p = 0.0004, adjusted p = 0.004) or of *limited value* (p < 0.0001, adjusted p < 0.0001). Taken together, these results suggest that PSA screening supporters are a group that, when confronted with evidence indicating that PSA screening prevents a much more limited number of deaths than they expect, their willingness to accept screening remains high, even in scenarios with minimal or no benefits. The file containing the results of the Chi Square tests is available on this study’s OSF repository [29].

#### PSA screening supporters generally have personal experience with PSA screening and cancer

We conducted an exploratory MANOVA to examine the relationship between demographic and health-related variables, and the two dependent variables identified before: the willingness to undergo PSA screening, and the perceived societal value of PSA screening. MANOVA results indicate some significant multivariate effects for education level (Roy’s greatest root, p = 0.0397), age (Roy’s greatest root, p = 0.0237), cancer diagnosis (Roy’s greatest root, p = 0.0316), and prostate cancer familiarity (Roy’s greatest root, p = 0.0148). The file containing the MANOVA is available on this study’s OSF repository [29]. We further explored the influence of these independent variables on the willingness to undergo PSA screening, and on the perceived societal value of PSA screening with ANOVA. As expected, individuals with a history of undergoing PSA screening were significantly more willing to undergo screening again (F = 8.33, p = 0.000002); participants with a history of cancer diagnosis were significantly more likely to show willingness to undergo PSA screening (F = 3.64, p = 0.0129); and familiarity with prostate cancer was also significantly associated with willingness to undergo screening (F = 2.85, p = 0.0236). For perceived societal value of PSA screening, the effect is similar: individuals with previous PSA screening experience perceived the societal value of screening as significantly higher than those without prior experience (F = 8.29, p = 0.000002); participants with a history of cancer were more likely to view PSA screening as having higher societal value (F = 3.38, p = 0.0184), as well as familiarity with prostate cancer (F = 3.26, p = 0.0120). Across both dependent variables, PSA screening history, previous cancer diagnoses, and familiarity with prostate cancer were the most consistent and significant predictors of participants’ willingness to undergo PSA screening and their perception of its societal value.

Additionally, participants who had never undergone PSA screening were significantly less willing to undergo screening compared to those who had been screened within the past year (mean difference = 0.62, p < 0.0001) and those who had been screened more than a year ago (mean difference = 0.39, p = 0.0218). This suggests that respondents who had a recent PSA screening experience are more likely to consider undergoing screening again. For perceived societal value of PSA screening, significant differences were observed in the post-hoc analysis: those who had never undergone PSA screening viewed PSA screening as significantly less valuable to society compared to those who had undergone screening in the past year (mean difference = 0.61, p < 0.0001) and those who had been screened more than a year ago (mean difference = 0.42, p = 0.0057). For cancer diagnosis, the post-hoc test highlighted a significant difference between participants with no previous cancer diagnosis and those previously diagnosed with prostate cancer. Participants with previously diagnosed prostate cancer were significantly more willing to undergo PSA screening (mean difference = 0.34, p = 0.0013), reinforcing the idea that personal health experiences drive willingness to undertake PSA screening. For the perceived societal value of PSA screening, there was a significant difference between participants who had no previous cancer diagnosis and those previously diagnosed with prostate cancer. Participants diagnosed with prostate cancer showed a higher perceived societal value of PSA screening (mean difference = 0.32, p = 0.0016). There was also a significant difference between participants previously diagnosed with prostate cancer and those previously diagnosed with other cancers (mean difference = -0.53, p = 0.0259), with prostate cancer patients perceiving PSA screening as more valuable. The file containing the ANOVAs and the Tukey tests is available on this study’s OSF repository [29].

These findings are supported by the thematic analysis of participants’ responses to the open-ended question, “Can you please explain why you answered yes?” This question was posed to those participants who indicated that PSA screening is beneficial, even in the hypothetical Scenario 0, where it was suggested that the screening would prevent no deaths. The most frequently cited reason, appearing 26 times, was due to their personal experience and family history, suggesting that a personal history of prostate cancer, either through direct or indirect experience or family history, heavily influenced their decision-making process.

Another prevalent theme, mentioned by 23 participants, was their belief in the importance of early detection and treatment. These participants valued the potential for early detection of cancer, even in the absence of a guaranteed life-saving outcome. Closely related, 17 participants mentioned peace of mind and knowledge as a significant motivator, reflecting a psychological benefit associated with knowing one’s health status. Furthermore, 13 participants mentioned the importance of prevention and proactive health management, even when the long-term benefits might be uncertain. Similarly, 12 participants indicated their willingness to balance the risks of overdiagnosis and overtreatment with the possibility of catching a serious condition early. Other participants mentioned that undergoing PSA screening is a routine and simple procedure, indicating that for some, participation in regular health screenings is not something problematic. The file containing the coded material is available on this study’s OSF repository [29].

## Discussion

Our study provides critical insights into public perceptions of PSA screening, revealing a significant divergence between participants’ understanding and expectations of PSA screening, and the established clinical evidence on the benefits of PSA screening. Notably, participants consistently overestimated the life-saving benefits of PSA screening, with many estimating that screening prevents hundreds of deaths per 1000 man screened, whereas the actual number is around 3 per 1000 men screened [7,11,12]. This overestimation persisted even after participants were provided with information and evidence about the risks of overdiagnosis and overtreatment, suggesting that beliefs and support for PSA screening may be driving their decisions more than evidence-based reasoning. It is important to clarify that here we are not claiming or suggesting that PSA screening holds no value, either individually or socially; rather, our findings highlight a significant mismatch between people’s perceptions and reality about current PSA screening technology benefits. This behavior suggests that for many individuals, undergoing PSA screening may offer psychological reassurance, irrespective of its clinical efficacy as a tool to prevent death from prostate cancer. The thematic analysis further underscores this point, reflecting a strong emotional or experiential attachment to PSA screening as a protective health measure, even when confronted with evidence suggesting benefits that are more limited than participants initially believed. The existence of these opinions about PSA screening also highlights an opportunity for public health authorities to reassess how they communicate the benefits and risks of PSA screening programs. Data suggest that public support for screening is not simply a matter of providing more or better information. Even in the face of clear evidence about the risks of overdiagnosis and limited benefits, many participants remain steadfast in their belief in the screening’s value. This belief system is akin to what is observed in other polarized health topics, where emotional or ideological beliefs take precedence over empirical data [30,31]. However, unlike vaccine skepticism, where the public rejects a health intervention, here, the public clings to a health intervention despite evidence that its benefits are less than they had initially perceived. The persistent support for PSA screening, even in unfavorable scenarios (see Scenario 0 in our survey), suggests that, for some people, commitment to PSA screening is not necessarily driven by a rational assessment of the risk-benefit ratio. Rather, it may be influenced by personal experiences, psychological benefits (e.g., reassurance and peace of mind), and general belief in the importance of early detection. This opens up an opportunity to implement screening programs that align more closely with public sentiment while also educating individuals about the actual risks involved; screening programs can be designed so that patients are fully informed about the potential harms and benefits of screening, without undermining the importance they place for their personal choices about health, leaving the patients in front of a fully autonomous decision.

Our exploration of preference epidemiology in the context of PSA screening suggests that this approach could be transferable to other screening programs, such as that of mammography [1–3], mentioned in the introduction, but also colonoscopy for colorectal cancer [32], or hypertension screening in children [33–35]. These programs similarly involve complex trade-offs between early detection and risks like overdiagnosis, false positives, and potential psychological impacts. By applying a preference epidemiology framework across different types of screenings, it could be possible to capture more accurately how people value these trade-offs, creating guidance that reflects both clinical evidence and patient perspectives. Improving health literacy through population-level communication strategies grounded in preference epidemiology could serve as a powerful intervention, helping individuals better understand the risk/benefit trade-offs involved in screening. While these surveys primarily aim to inform public health guidance and align screening programs with broader population sentiment, they can also positively impact individual decision-making, as by encouraging individuals to reflect on specific trade-offs, they can support more informed, autonomous decisions about whether to undertake screening, enhancing their ability to interpret results and understand how screening outcomes could influence their lives. Communication approaches based on evidence collected with preference epidemiology studies could potentially serve as an effective intervention for improving health literacy, helping individuals to better understand the risk/benefit trade-offs involved in screening and allowing them to make more informed and therefore more autonomous decisions about whether to undertake screening – being able to interpret the results, and to understand how the screening can influence their lives. In light of these findings, we suggest that public health strategies should not only focus on disseminating accurate information in the context of PSA screening but also consider the emotional and experiential factors that influence decision-making and that may arise in response to overtreatment and overdiagnosis. Addressing public beliefs through tailored communication that resonates with individuals’ values and experiences, rather than solely relying on factual information, may be more effective in guiding decision-making about PSA screening towards a more informed approach.

### Limitations

Despite the promising insights, our study has limitations. First, recruitment via Facebook ads has likely led to an overrepresentation in our survey of men interested in PSA screening and prostate cancer. Additionally, the reliance on self-reported data may have introduced response bias, as participants might have misremembered their experiences. Finally, the study did neither consider nor analyse the influence of exposure to media narratives or public health campaigns, that could have influenced participants’ responses.

## Conclusion

This study reveals a significant gap between public perceptions of PSA screening and the actual clinical evidence of the benefits of PSA screening. Participants consistently overestimated the benefits of screening, with many maintaining their support even when presented with the information related to the risks of overdiagnosis and the actual number of deaths prevented. This suggests that decision-making regarding PSA screening is not solely driven by evidence-based considerations but can be driven by the existence of polarized opinions about the screening itself due to personal experiences with prostate cancer and PSA screening. In this case people tend to maintain their unconditional support for PSA screening measures despite evidence suggesting its benefits are considerably lower than they initially believed. This presents a challenge for public health communication: simply providing factual information about PSA screening may not be sufficient to help individuals reach fully autonomous decisions about whether to undertake PSA screening in light of its benefit/risk profile. This is where preference epidemiology studies can offer valuable support, as they allow to explore how individuals weigh the benefits and risks of medical interventions such as PSA screening, taking into account their values and preferences. By understanding these decision-making processes, we can design more personalized and effective public health strategies that resonate with the public, ultimately guiding individuals toward informed decisions that resonate with their personal values.

## Data Availability

All the data are available on this study's OSF repository - DOI: 10.17605/OSF.IO/BFY7T.

https://osf.io/bfy7t/

## Authors’ declarations

### Ethics approval

The study was reviewed and approved by the Institutional Review Board (IRB) of the Faculty of Medicine, University of Zurich (reference number 2024-255652).

### COI

The authors declare that there are no conflicts of interest related to this study.

## Notes

### Competing Interest Statement

The authors have declared no competing interest.

### Funding Statement

This study was funded by UZH DSI - Digital Society Initiative, Mind the Patient lab.

